# Hyperglycemia in pregnancy: A sensitivity analysis study of two recommended screening tests in Tanzania

**DOI:** 10.1101/2025.05.08.25327275

**Authors:** Amani Kikula, Kaushik Ramaiya, Nathanael Sirili, Matilda Mkonyi, Rukia Msumi, John Macha, Andrea B. Pembe, José L. Peñalvo, Lenka Beňová

**Affiliations:** Department of Obstetrics and Gynaecology, Muhimbili University of Health and Allied Sciences, Dar es Salaam, Tanzania; Department of Family Medicine and Population Health, University of Antwerp, Antwerp, Belgium; Department of Public Health, Institute of Tropical Medicine, Antwerp, Belgium; Department of Internal Medicine, Shree Hindu Mandal Hospital, Dar es salaam, Tanzania; Department of Development Studies, Muhimbili University of Health and Allied Sciences, Dar es Salaam, Tanzania; Department of Obstetrics and Gynaecology, Mbagala Rangi Tatu Hospital, Dar es salaam, Tanzania; Department of Obstetrics and Gynaecology, Kisarawe District Hospital, Coast, Tanzania; Instituto de Salud Carlos III, Madrid, Spain

**Keywords:** Gestational diabetes mellitus, screening, sensitivity, pregnancy, Tanzania, non-communicable disease

## Abstract

Screening for hyperglycaemia in pregnancy (HIP) is an entry point to a healthy pregnancy and childbirth for both the woman and child, and offers a window of opportunity for the prevention of cardio-metabolic complications over the life-course of the woman and newborn. In Tanzania, two different screening algorithms to identify women with HIP are recommended, with no clear understanding of which performs better. We aimed to determine the prevalence of hyperglycemia in pregnancy (HIP) among antenatal care (ANC) attendees and the accuracy of HIP screening tests.

A cross-sectional study design involving 970 women attending ANC clinic was done in two district hospitals (Mbagala Rangi Tatu and Kisarawe) of Tanzania between June and October 2024. Socio-demographic, obstetric characteristics, and anthropometric parameters were obtained from study participants. A checklist screening test (CST) for gestational diabetes mellitus (GDM), defined by the Tanzanian standard treatment guideline, was used. We tested for glycosuria and two-hour 75-gram oral glucose tolerance test (OGTT) on every study participant. We conducted descriptive statistics and sensitivity analysis to determine the prevalence of HIP and assess the screening performance of CST and glycosuria test against OGTT using World Health Organization 2013 diagnostic criteria for GDM.

The prevalence of HIP was 10% (7.9% GDM and 2.1% diabetes in pregnancy). Glycosuria test missed all women with GDM and could identify only 20% with diabetes in pregnancy. The CST had a sensitivity and specificity of 72.2% and 32.4%, respectively, while the glycosuria test had 4.1% and 97.1%. The area under the receiver operator characteristic curve for both the CST 0.523 (0.48-0.57,95%CI) and glycosuria test 0.506 (0.49-0.53,95%CI) was low, indicating their poor discriminatory performance for HIP.

One in every ten women had HIP. The CST is a better first-step screening test than the glycosuria test based on sensitivity.

## 1 Introduction

In 2020 , approximately 800 women died globally every day due to pregnancy-related complications with 70% of these deaths occurred in sub-Saharan Africa (SSA) (1). Complicating the efforts to reduce maternal deaths is the increasing contribution of indirect causes to overall maternal mortality, known as the obstetric transition (2–4). It is in part caused by the growing epidemic of overweight and obesity, particularly among women of SSA (5–7). In Tanzania, like in most other SSA countries, the prevalence of high body mass index (BMI) among adults is increasing (8,9) and over a quarter of women who intend to conceive were either overweight or obese (10).

Overweight and obesity during pregnancy are risk factors for gestational diabetes mellitus (GDM) (11). GDM increases the risk for complications during pregnancy, childbirth, and beyond, including pre-eclampsia and intra-uterine fetal death; traumatic or operative delivery (12). Following delivery, the risk for infection and thromboembolism is higher among women with GDM (12). The risk for developing type 2 diabetes mellitus and other cardiometabolic conditions later in life is also higher for the newborn and woman who had unmanaged GDM (13). Thus, screening for GDM as an entry point to successful timely management of the condition is an important predictor of a healthy pregnancy and childbirth for both the woman and child, and offers a window of opportunity for the prevention of these complications over the life-course (14).

In 2013, the World Health Organization (WHO) adopted the International Association of Diabetes and Pregnancy Study Groups recommendation for universal screening using a one-step, 75-grams, two-hour oral glucose tolerance test (OGTT) (15). Alternatively, a two-step approach involves an initial screening based on one or more risk factors to identify women who require an OGTT. This method is favored in settings where universal OGTT-based screening is not economically feasible. The International Federation of Gynecology and Obstetrics (FIGO) advocates for context-specific screening strategies that align with the healthcare system realities of different countries (12).

In Tanzania, the two-step approach has been adopted. However, two different screening algorithms to identify women for the second step co-exist in national guidelines. The Tanzania antenatal care (ANC) guideline uses glycosuria as the initial screening step for those who would require a random blood glucose test (16). The Tanzania standard treatment guideline (STG) relies on a checklist of risk factors (Table 1) to identify those who should be further tested using OGTT (17). In the Tanzanian context, there is no evidence about which one of these screening modalities performs better. Therefore, this study aimed to assess the prevalence of GDM and evaluate the accuracy of both screening algorithms among women attending ANC in two district hospitals in Tanzania.

**Table 1:** Source and information obtained from the woman during data collection.

| Source | Variable |
| --- | --- |
| Extracted from the woman-held ANC record | Age, place of residence, education level, gravidity, parity, HIV status, last normal menstrual period, body weight at first ANC visit |
| Interview | Checklist screening variables: Body mass index (BMI), history of GDM, previous history of delivery of a baby weighing >4kg at birth, history of stillbirth or early neonatal death, family history of diabetes mellitus, grand multiparity, Additional variable: marital status. |
| Measured | Weight, height, glycosuria<sup>a</sup>, mid-upper arm circumference, blood glucose |

## 2 Materials and methods

We conducted a cross-sectional study to assess screening accuracy for hyperglycemia in pregnancy (HIP) with a focus on sensitivity of the screening tests (18). The index screening tests were a checklist screening test (CST) and a urine dipstick glycosuria test. The diagnostic gold standard test was the two-hour 75-gram non-fasting OGTT.

### 2.1 Study sites

We collected data in ANC clinics of two primary-level district hospitals in Tanzania between June 19 and October 31, 2024. Firstly, Kisarawe hospital is located in a rural district of the Coast region and Mbagala Rangi Tatu hospital is in an urban district of Dar es salaam region. More details about study sites are provided elsewhere (19). In Tanzania, ANC service is offered from the dispensary level to the national level hospital, with 90% of women receiving ANC services from skilled providers (20). More women (55%) are receiving ANC at district hospitals compared to any other level (21).

### 2.2 ANC services related to GDM care

During ANC, women’s height is measured at the first visit; their weight and blood pressure are measured every visit. The two study hospitals had laboratory departments that served the whole hospital, including the ANC clinic. Routinely, point-of-care capillary blood glucose tests to check for glucose levels were used and either a urine dipstick or analyzer machine for urinalysis was done. All care provided and any results were documented in the hospital registers and the woman-held ANC record. Services provided at the ANC clinic are guided by the Tanzanian ANC guideline (16) and supplemented by the Tanzanian STG (17).

### 2.3 Sample size and eligibility

We used a standard sample size formula for sensitivity studies (22). A minimum sample size of 946 women was deemed necessary considering a 19.5% prevalence of GDM (23) and expected level of sensitivity for glycosuria test of 4.7% and 69% for CST (24,25). The desired precision estimate of 7% at a 95% level of confidence, and a 10% refusal rate.

All women attending ANC clinics in the study hospitals at gestation age of 24 to 28 weeks were approached and those who provided informed written consent were enrolled for a structured interview and other measurements (Table 1). We excluded women who were known diabetes mellitus patients through medical records or self-report, who were on steroid treatment for any medical condition, or who had a self-reported active diagnosed infectious disease (e.g. malaria) on day of the visit. We consecutively included all eligible women in both hospitals until the sample size was reached.

For the glycosuria test, women were provided with urine collecting containers (60ml) and instructed to collect a minimum of 10ml of urine within dedicated hospital toilets. They immediately submitted the urine sample to the trained laboratory research assistants (RAs) for processing.

Following collection of the urine sample, study participants were provided with a rapillose 75gram OGTT solution (26) and we supervised their consumption in 15-20 minutes. RAs documented the participants’ study ID and time of intake of the OGTT solution in the laboratory study request form and attached it to the woman’s ANC record. Women then received their routine ANC while awaiting the two-hour time mark for a blood glucose test. Fig 1 shows the sequence of study procedures that recruited women in the study followed. Laboratory RAs were selected laboratory personnel from each hospital who received and processed the urine for glycosuria and POC capillary blood glucose tests. Calibration was done after every 20^th^ test using calibration strips and a control solution provided by the manufacturer.

**Fig 1:**
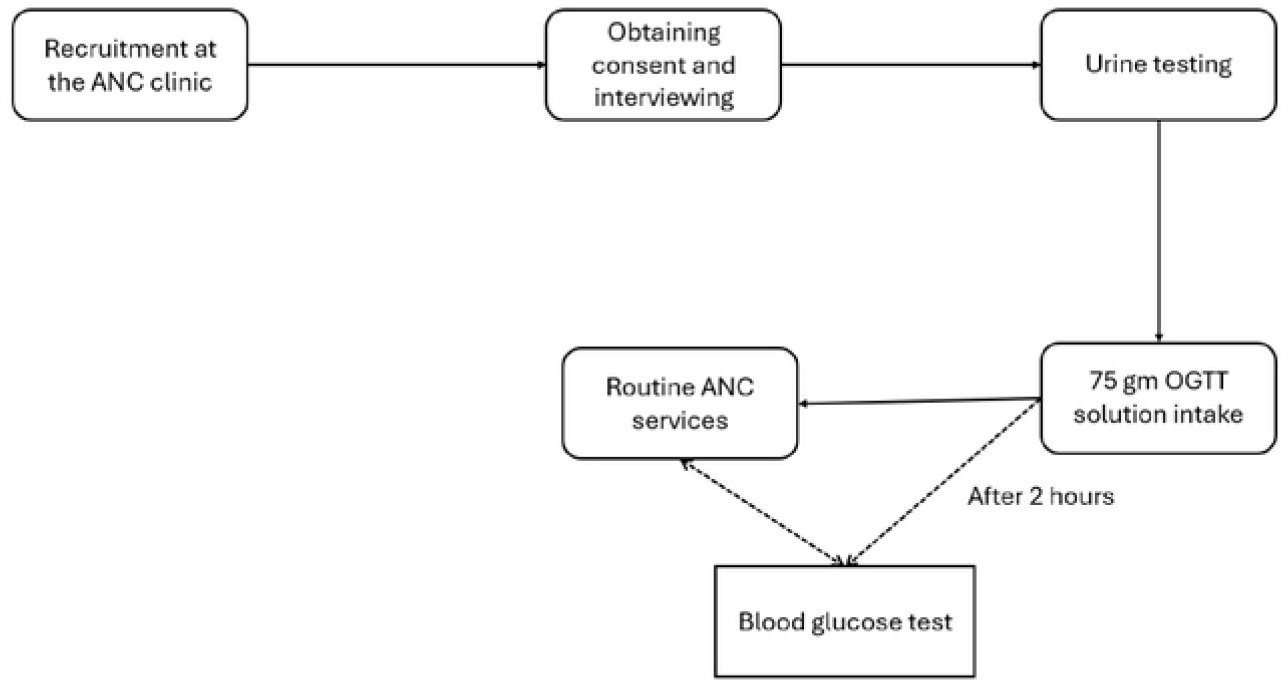
Description of recruitment of study participants and study procedures

The results of the glycosuria test and the blood glucose test were documented in the study request form and the ANC record. A member of the research team also communicated the test results to the woman and the ANC clinic staff.

Women with blood glucose of ≥8.5 mmol/dl were referred to the hospital obstetrician and continued with further care as per hospital protocol. Table 2 describes all the measured parameters that were done to recruited women.

**Table 2:**
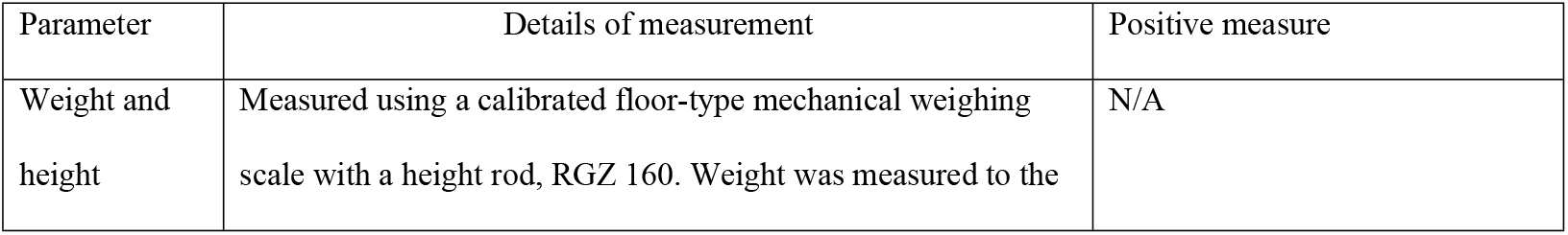
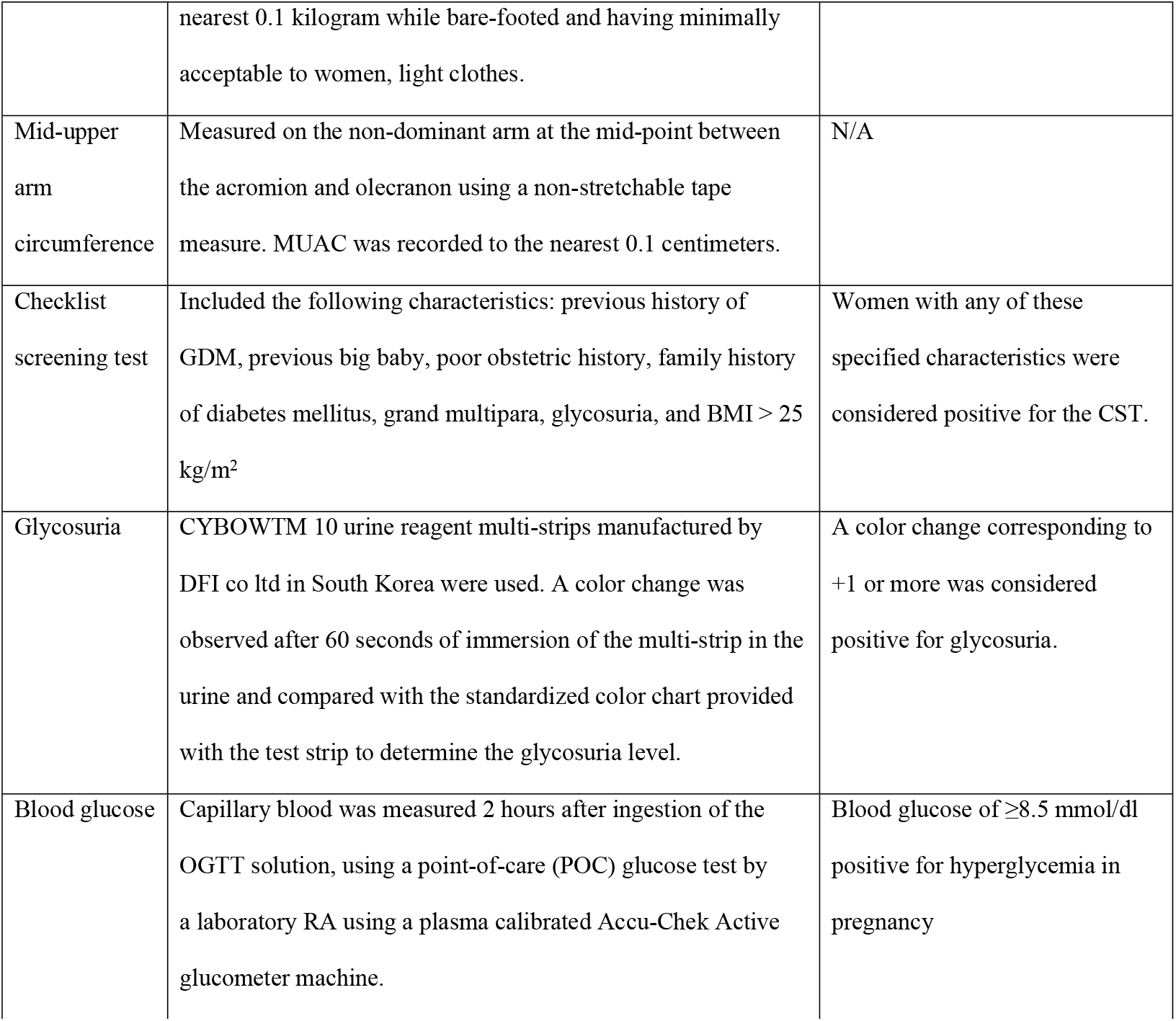
Description of the measured parameters for study participants.

### 2.4 Data collection and analysis

We collected all the data using kobo toolbox electronic software, extracted and cleaned in Excel spreadsheet then exported for analysis into STATA SE software (V.18.5). All entries with a missing blood glucose value were excluded from the analysis.

We conducted pooled descriptive and sensitivity analysis of participant characteristics from both hospitals. For each screening test and the diagnostic test, we categorized results as either positive or negative.

The BMI was extrapolated from the MUAC with a consideration that MUAC has a negligible increase during pregnancy (0.05 cm) and a MUAC of ≥ 28 cm was considered to correlate to pre-pregnancy BMI of ≥ 25kg/m^2^ (23,27). In Tanzania, only 34% of women have their first ANC clinic visit in the first trimester (20), this limited using the documented body weight on the ANC record for BMI calculation.

We used a ≥8.5mmol/L cut-off value for the diagncostic test to determine the true positive, false positive, true negative, and false negative values. The performance of the screening tests was measured using sensitivity, specificity, negative and positive predictive values, with a 95% confidence interval (CI). Hyperglycemia in pregnancy includes GDM and diabetes in pregnancy (DIP). We classified blood glucose levels of 8.5-11.0 mmol/L as GDM, and >11.0 mmol/L as DIP (12).

We compared the performance of the two screening tests using the receiver operator characteristics (ROC) curves against the diagnostic OGTT test. The diagnostic test result was dichotomized to hyperglycemia and normal blood glucose value (<8.5 mmol/L).

### 2.5 Ethical approvals

The study received ethical approval from the Institutional Ethics Committee at the Institute of Tropical Medicine, Antwerp 1687/23, Muhimbili University of Health and Allied Sciences Research and Ethics committee MUHAS-REC-07-2023-1834 and the National Health Research Ethics Review Committee NIMR/HQ/R.8a/Vol.IX/4457.

## 3. Results

We recruited a total of 975 women for screening at the two hospitals. Out of these, five were excluded; two were unavailable for the two-hour blood glucose test, and three requested withdrawal from the study (Fig 2).

**Fig 2:**
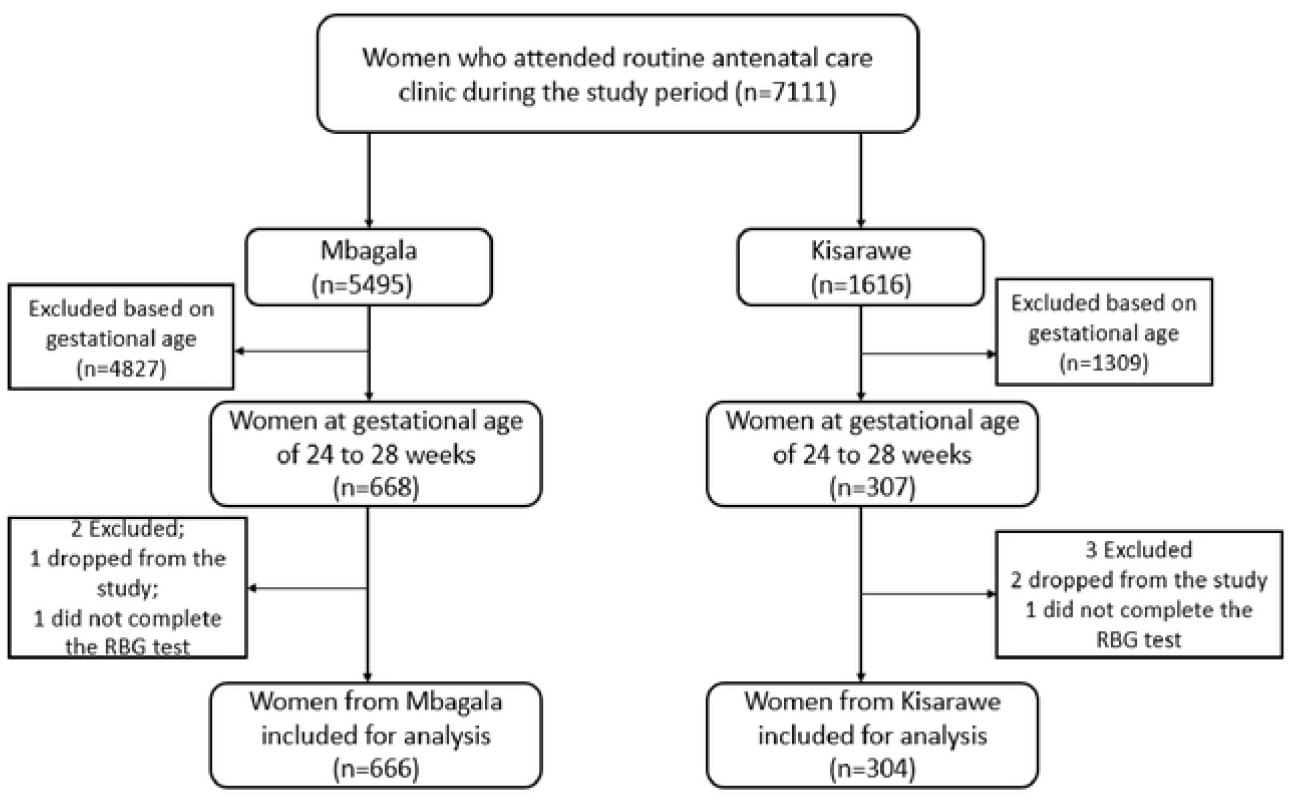
Study flowchart for inclusion of study participants for analysis

Out of the 970 women included in the final analysis sample, 68.7% (666 women) were recruited at Mbagala Rangi Tatu Hospital (Table 3). Just half (52.2%) of the study participants were in the 20–29-years age group. A total of 40.2% had attained secondary school-level education and the majority (91.6%) were either married or living with a partner. About two-thirds (56.4%) had a BMI of ≥25kg/m^2^ and 4.1% were HIV-positive.

**Table 3:** Sociodemographic characteristics, HIV status, and body mass index of the study participants by hospital (n=970)

| Characteristic | Hospitals |  |  |
| --- | --- | --- | --- |
|  | Mbagala Rangi |  |  |
|  | Tatu<br>(n=666) | Kisarawe<br>(n=304) | Total<br>(n=970) |
| **Age group** | n (%) | n (%) | n (%) |
| <20 | 55 (8.2) | 23 (7.6) | 78 (8.0) |
| 20-29 | 341 (51.2) | 165 (54.2) | 506 (52.2) |
| 30-39 | 243 (36.5) | 103 (33.9) | 346 (35.7) |
| ≥40 | 27 (4.1) | 13 (4.3) | 40 (4.1) |
| **Highest level of education** |  |  |  |
| No formal education | 6 (0.9) | 14 (4.6) | 20 (2.1) |
| Until primary school | 339 (50.9) | 187 (61.5) | 526 (54.2) |
| Secondary school | 301 (45.2) | 89 (29.3) | 390 (40.2) |
| College/University | 20 (3.0) | 14 (4.6) | 34 (3.5) |
| **Married/Living with a partner** |  |  |  |
| Yes | 606 (91.0) | 282 (92.8) | 888 (91.6) |
| No | 60 (9.0) | 22 (7.2) | 82 (8.4) |
| **Parity** |  |  |  |
| 0 | 210 (31.5) | 103 (33.9) | 313 (32.3) |
| 1 | 170 (25.5) | 81 (26.6) | 251 (25.9) |
| 2 - 4 | 276 (41.5) | 117 (38.5) | 393 40.5) |
| ≥5 | 10 (1.5) | 3 (1.0) | 13 (1.3) |
| **MUAC-adjusted BMI** |  |  |  |
| <25 | 227 (34.1) | 196 (64.5) | 423 (43.6) |
| ≥25 | 439 (65.9) | 108 (35.5) | 547 (56.4) |
| **HIV status** |  |  |  |
| Positive | 33 (5.0) | 6 (2.0) | 39 (4.1) |
| Negative | 625 (93.8) | 292 (96.0) | 917 (94.5) |
| Missing | 8 (1.2) | 6 (2.0) | 14 (1.4) |

The prevalence of GDM was 7.9%, and of DIP 2.1%, with a combined 10.0% prevalence of HIP. The glycosuria test was negative for all women diagnosed with GDM and identified only 20% of women with DIP. The CST identified over two-thirds (67.5%) of women with GDM and 90.0% of women with DIP (Table 4).

**Table 4:** Prevalence of gestational diabetes mellitus and diabetes in pregnancy among women from the study hospitals (n=970)

| Characteristic |  |  |  |
| --- | --- | --- | --- |
|  | Normal, n (%) | Gestational diabetes mellitus, n (%) | Diabetes in pregnancy, n (%) |
| **Diagnostic test** |  |  |  |
| 2 hour OGTT | 873 (90.0) | 77 (7.9) | 20 (2.1) |
| **Screening test** |  |  |  |
| **Glycosuria** |  |  |  |
| Positive | 25 (2.9) | 0 (0.0) | 4 (20.0) |
| Negative | 848 (97.1) | 77 (100.0) | 16 (80.0) |
| **Checklist** |  |  |  |
| Positive | 616 (70.6) | 52 (67.5) | 18 (90.0) |
| Negative | 257 (29.7) | 25 (32.5) | 2 (10.0) |
| **Hospital** |  |  |  |
| Mbagala Rangi Tatu | 595 (89.3) | 56 (8.4) | 15 (2.3) |
| Kisarawe | 287 (94.7) | 21 (6.9) | 5 (1.6) |

The CST had a higher sensitivity than glycosuria test (72.2% vs 4.1%), making the CST a better first-step screening test as compared to the glycosuria test. The glycosuria test had a better specificity of 97.1% than 32.4% of the CST. Both tests had comparable positive predictive values (10.6% CST,13.8% glycosuria) and negative predictive values (91.3% checklist and 90.1% glycosuria) (Table 5). Overall, both tests performed poorly as diagnostic tests for HIP with area under the ROCs of 0.52 (0.48-0.57, 95%CI) and 0.51 (0.49-0.53, 95%CI) respectively, indicating poor discrimination (Figure 3).

**Table 5:**
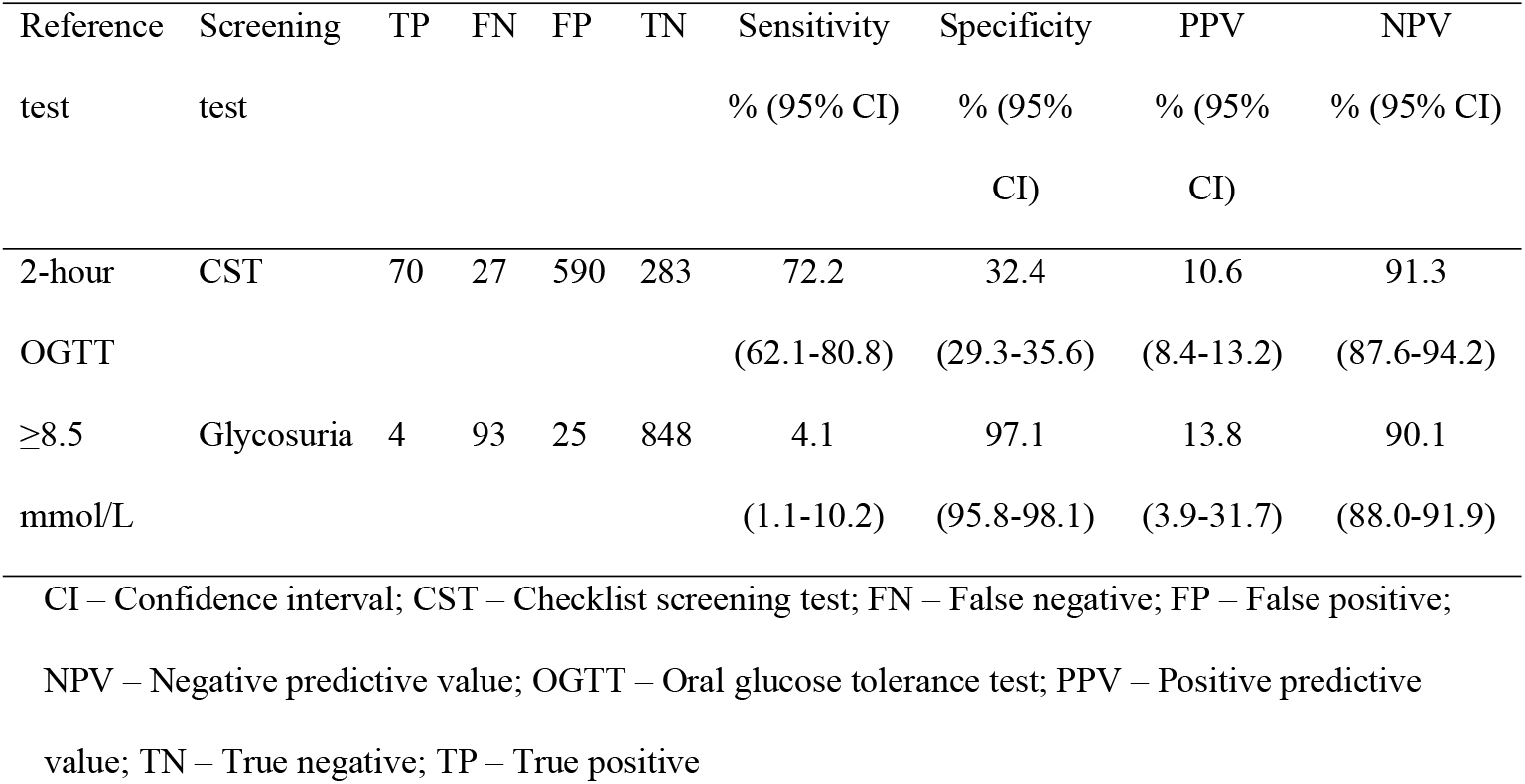
Sensitivity analysis for Hyperglycemia in pregnancy of checklist screening test and glycosuria test against 2-hours OGTT (n=970)

**Fig 3:**
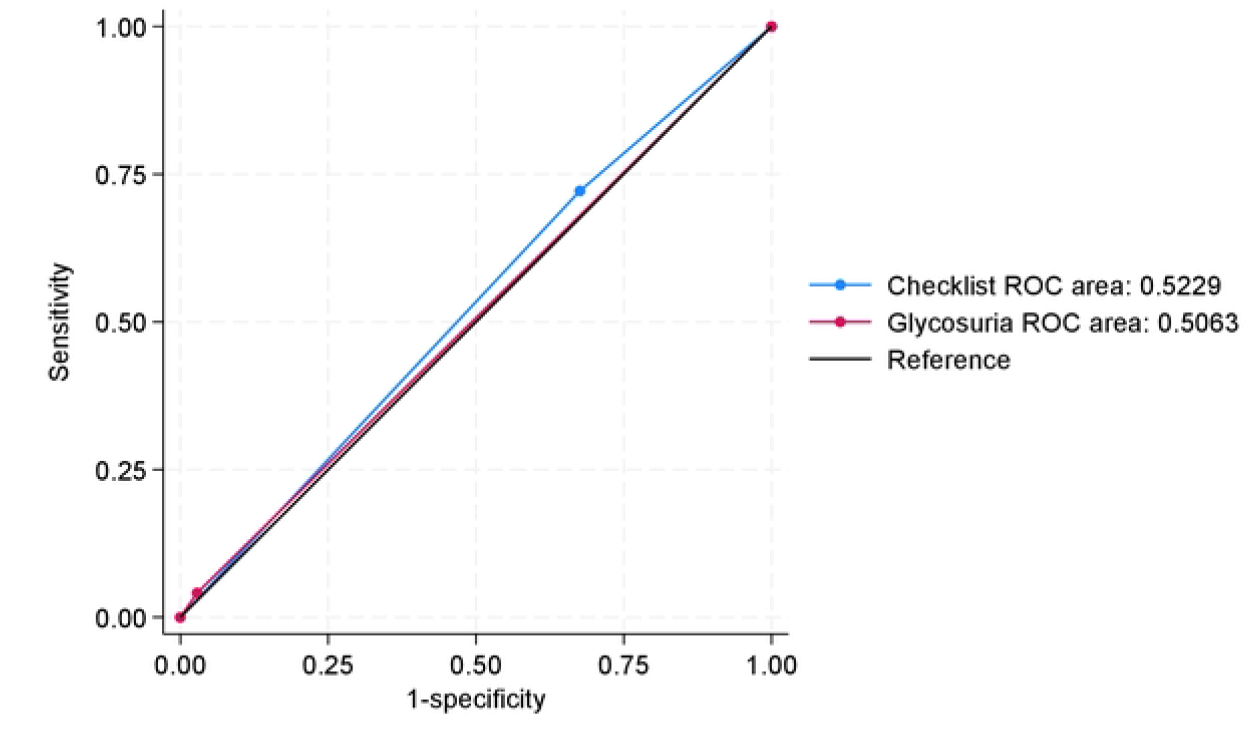
Receiver observer characteristics curve comparing checklist screening test and glycosuria tests for Hyperglycemia in pregnancy.

## 4 Discussion

In the two study hospitals, one in every ten women presenting for antenatal care at 24 to 28 weeks of pregnancy had hyperglycemia in pregnancy. The glycosuria test could not identify any woman with GDM and missed more than three-quarters of those who tested positive for DIP. The CST had better sensitivity than the glycosuria test making it a better first-step screening test than the glycosuria test in identifying women with hyperglycemia during pregnancy for diagnostic tests. Both CST and glycosuria tests cannot be used as stand-alone diagnostic tests for Hyperglycemia in pregnancy.

At a global level, multiple guidelines with different algorithms exist for GDM screening (28). Screening algorithms and diagnostic thresholds differ across different available guidelines and have evolved affecting GDM prevalence estimation and its standardization (29–32). The FIGO recommended standardized algorithms and tests for GDM screening to suit different contexts, including having the OGTT as the gold standard test for GDM diagnosis, however, its cost has limited its universal usage, particularly in low-resource countries (12,33). Tanzania faces a unique situation of having two co-existing algorithms with a glycosuria test and CST recommended independently for GDM screening in the two available national guidelines (16,17). The healthcare workers using these guidelines work within a health system with limited supplies, training opportunities, and monitoring frameworks (21). These challenges are known to limit the implementation of multiple clinical guideline recommendations (34) with low levels of optimal adherence to available treatment guidelines (35–37). Having two guidelines in Tanzania, with two algorithms and different types of tests for screening, further complicates efforts to mainstream GDM care within the ANC setting. This underscores the need for the Ministry of Health to have a single algorithm for GDM screening in its guidelines. This should go hand-in-hand with efforts to improve the health system capacity to manage those who will test positive for GDM (38).

For the study participants, the glycosuria test missed all women who screened positive for GDM and was positive for only one-fifth of those with DIP. The urine multi-stick used for glycosuria test is routinely used in most primary healthcare facilities for assessing other clinical parameters in urine as part of ANC check (20), however, it lacks the required sensitivity as a GDM first-step screening test.

The CST demonstrated better sensitivity compared to glycosuria test. The items on the checklist could be further adapted to the Tanzanian context to improve its sensitivity and specificity as a first-step screening test (24,39). Studies have reported that using additional characteristics such as body fat content, history of macrosomic birth, MUAC, and family history of diabetes improved the CST’s screening accuracy (39). However, it requires a good medical records system to report such characteristics (40). Given the low sensitivity of the glycosuria test shown in this study and other studies (12,25), it should no longer be used as a GDM screening tool in Tanzania.

Globally, about one in every ten pregnancies are affected by GDM, (41) in line with the findings from this study. In North Africa, over a quarter of pregnant women have GDM, which is the highest in Africa (42). Overall, there is no significant difference in the prevalence of GDM between low-income and high-income countries (42). In Tanzania, while there was no GDM detected in the early 1990s (43), GDM prevalence since the 2000s have ranged from 4.3% to 39% (23,44,45). These variations could be attributed to differences in the studies’ cut-off values and tests used for diagnosis, the population studied, but are also sounding an alarm about the rise of GDM in the country. Without routine screening during ANC, pregnant women will miss the opportunity for early detection and timely intervention to prevent potential GDM-related complications during pregnancy and cardiometabolic complications after childbirth (46). The high prevalence and known morbidities that result from GDM underscore the need for universal screening of GDM to achieve targeted reductions in maternal mortality in the global south.

We acknowledge using a non-fasting two-hour OGTT as a diagnostic test for GDM which is evidenced to have similar diagnostic accuracy to the fasting two-hour OGTT (12,47,48). The blood sample used was a capillary blood sample which has slight lower glucose levels to the plasma blood samples. Despite these limitations, to our knowledge, this is the first study of its kind in Tanzania to compare the screening potential of two nationally recommended screening tests for GDM. Additionally, our use of standardized 75-gram OGTT solutions maintained consistent glucose loads to all participants. We believe that these findings highlight the need to urgently revise the GDM screening guidelines and consistently implement one well-performing screening algorithm.

## 5 Conclusions

Hyperglycemia in pregnancy was found in 10% of the pregnant women in the study hospitals. The CST performed better as a first-step screening tool. Glycosuria test performed poorly and should not be used as a screening tool for GDM. We recommend further studies to be done to inform the revision of the GDM screening tests to have one with an optimally acceptable discriminatory potential during screening for further diagnostic confirmation of hyperglycemia in pregnancy in Tanzania.

## Data Availability

Data will be available upon reasonable request through the National Institute of Medical Research, Tanzania - National Health Research Ethics Review Sub-committee - www.nimr.or.tz

## Acknowledgments

We thank the women who agreed to participate in this study. We also acknowledge Judith Banyenza, Karama Ogillo and Joyce Kaswamila who assisted in data collection, Davis Mlay for managing the electronic database. We are also thankful to the health care workers and health managers in the two districts that the work was done.

## Authors’ contributions

Conceptualization, data curation, formal analysis, investigation, methodology, project administration, resources, validation, visualization, writing original draft and reviews – AK; investigation, methodology, reviewing and editing the manuscript – MM, RM, and JM; conceptualization, supervision, methodology, review and editing of manuscript, and validation KR,NS, ABP, JLP, and LB.

